# Clinical Virology and Effect of Vaccination and Monoclonal Antibodies against SARS-CoV-2 Omicron Sub Variant BF.7 (BA.5.2.1.7): A systematic review

**DOI:** 10.1101/2022.12.25.22283940

**Authors:** Santenna Chenchula, Krishna Chaitanya Amerneni, Mohan Krishna Ghanta, R Padmavathi, Madhu Bhargavi Chandra, Madhu Babu Adusumilli, Sofia Mudda, Madhavrao Chavan, Rupesh Gupta, Bhawna Lakhawat

**Affiliations:** Department of Pharmacology, All India Institute of Medical Sciences (AIIMS)Bhopal, Madhya Pradesh, India; Faculty, Western Michigan University, Kalamazoo, Michigan; Department of Pharmacology, MVJ Medical College and Research Hospital, Bangalore, Karnataka; SVS Medical College and Hospital, Telangana, India; All India Institute of Medical sciences, Bhopal; Department of Pharmacology, All India Institute of Medical Sciences (AIIMS) Mangalagiri, Andhra Pradesh, India; Department of Internal Medicine, Government Medical College, Shahdol, Madhyapradesh, India; department of Pharmacology, All India Institute of Medical sciences, Bhopal

## Abstract

Since its identification, the novel coronavirus “severe acute respiratory syndrome coronavirus 2 “(SARS-CoV-2) In in late 2019 AT Wuhan, China, by the World Health Organization (WHO), which cause the coronavirus disease 2019, is rapidly spreading, resulting in the global pandemic. As of 19 December 2022, more than 64 million confirmed cases and 6,645,812 deaths have been reported across the world. Over time, the SARS-CoV-2 acquired genetic mutations resulting in multiple types of SARS-CoV-2 variants and subvariants that have been confirmed. The Omicron (B.1.1.529) variant was identified later in November 2021, with enhanced immune escape and was followed with various sublineages due to mutations in the spike protein of the SARS-CoV-2. However, rapid resurge in COVID-19 reports by Omicron subvariant BF.7(BA.2.75.2) in China and other countries, alarming global threat. The present systematic review was conducted using the MeSH terms and keywords “Omicron” AND “BA.5.2.1.7” OR “BF.7” in Pub Med, Google Scholar and MedRXiv database and grey literature from the authentic database and websites. We identified a total of 14 eligible studies. We have reviewed all the eligible available studies to understand the viral mutations, and factors associated with the increase in the reports of COVID-19 cases in China and across the world and to evaluate the effectiveness of vaccination and monoclonal antibodies against the BF.7 variant.

## Introduction

Since its identification in late 2019 the novel coronavirus, severe acute respiratory syndrome coronavirus 2 (SARS-CoV-2) In Wuhan, China, by the World Health Organization (WHO), which cause the coronavirus disease 2019, it is rapidly spreading, resulting in the global pandemic [1]. As of 19 December 2022, more than 64 million confirmed cases and 6,645,812 deaths have been reported across the world [2]. Over time, the SARS-CoV-2 acquired genetic mutations resulting in multiple types of SARS-CoV-2 variants and subvariants that have been confirmed [3]. Certain variants have gained keen attention because of their characteristics of rapid transmissibility, enhanced immune escape and severity of the infection and these are considered variants of concern (VOC) that continue to threaten public health [3].

Later in November 2021, the Omicron (B.1.1.529) variant with enhanced immune escape was first reported from Botswana and thereafter from South Africa with an increased infection [3-4]. Very soon it has been spread swiftly to several other countries, across the world with subtle raise in the number of COVID-19 infections [4-5]. Subsequently, Omicron sublineages with increasingly greater replication advantages emerged, replacing the previous predominant sub lineage [3]. There are about 200 sublineages of Omicron [3]. The original Omicron variant was sublineage BA.1, BA.2, BA.3, BA.4 and BA.5 [5-6]. Other Omicron sublineages, such as BQ.1, BQ.11, BF.7, BA.2.75, and XBB, which evolved from various previously circulating sublineages, have been increasing in prevalence worldwide [7]. Each sublineage differs from the others by several mutations in the spike protein except for BA.4 and BA.5, which have identical spike proteins [8]. Evidence has shown that all omicron sub-variants are distinct from pre-omicron variants, including BA.1, BA.2 and BA.5 omicron sub-variants are also antigenically distinct from each other [9]. The BF.7 also known as BA.2.75.2 Variant was first identified on May 13, 2022, in Belgium. In response to the current surge in the COVID-19 reports by Omicron subvariant BF.7 also known as BA.2.75.2 in China and other countries, triggering global alarm the present review was conducted to understand the virology, factors associated with increased reports of COVID-19 infections with BF.7 variant in China and possible urgent preventing strategies to be taken to curtail the novel omicron variants outbreak across the world.

## Methodology

We have performed a comprehensive literature search from inception to till December 2022, using the MeSH terms and keywords “Omicron” AND “BA.5.2.1.7” OR “BF.7” in Pub Med, Google Scholar and MedRXiv database and grey literature from the authentic database and websites to find relative information about Omicron subvariant BF.7 (BA.5.2.1.7) and data assessed to understand viral mutations, factors associated with increased reports of COVID-19 infections with BF.7 variant in China and on literature evidence on vaccine effectiveness and monoclonal antibodies(mAbs) against BF.7 to understand possible urgent preventing strategies to be taken to curtail the novel omicron variants outbreak in China and across the world.

## Results

In the present review, after removing duplications and literature not mentioned regarding BF.7 or BA.5.2.1.7, a total of 14 studies were found eligible among the 79 studies **[Figure1]**. The SARS-CoV-2 Omicron BF.7 (BA.5.2.1.7) a subvariant of BA.5, is responsible for the ongoing resurgence of cases since late September 2022 in China [10].To date, BF.7 (BA.5.2.1.7) subvariant has been reported in Belgium, China, Denmark, Norway, France, Germany, India, Mongolia, the United Kingdom and the United States[11-12]. However, Before China, the BF.7 variant has been circulating at an incredibly intense level in USA and Europe since August 2022[8-10]. The current reports from China indicate BF.7 has the strongest infection ability out of the Omicron subvariants, being quicker to transmit than other variants, having a shorter incubation period, and with a greater capacity to infect people who have had a previous COVID infection, or been vaccinated, or both [11]. The most common symptoms of an infection with BF.7 were similar to those associated with other Omicron subvariants, primarily upper respiratory symptoms [12]. Patients may have a fever, cough, sore throat, running nose and fatigue, and gastrointestinal symptoms like vomiting and diarrhoea among other symptoms [11-12]. The basic reproduction number *Rt* of omicron BF.7(BA.5.2.1.7) variant was 10 to 18.6. which is significantly high compared to BA.1 Omicron variant which is only 5.08[11]. A study by Kathy Leung et al., to track the effective reproduction number *Rt* of Omicron BF.7 in Beijing from November to December 2022 found that the *Rt* increased to 3.42 (95% CrI: 2.79 – 4.17) on

**Figure 1.**
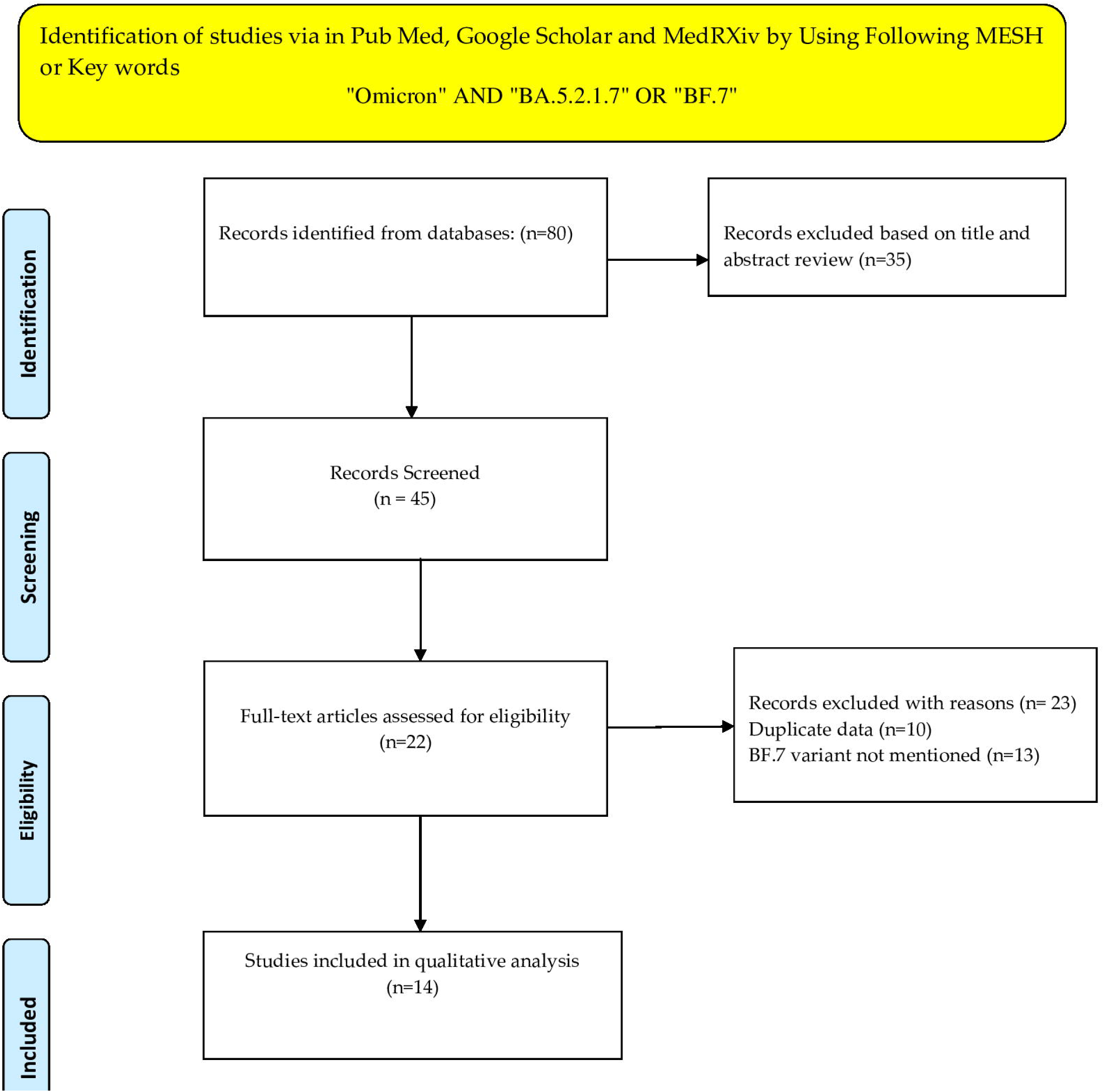
PRISMA Flow Diagram for the study selection.

November 18, the infection incidence peaked on December 10, and the cumulative infection attack rate was 42.5% (95% CrI: 20.3 – 63.9) on December 14[13]. The high transmission rate of BF.7(BA.5.2.1.7), is due to the novel mutations in the spike protein [11-15]. Evidence from the molecular modelling studies revealed the mechanisms of antibody-mediated immune evasion by R346T, K444T, F486S, and D1199N mutations on the receptor-binding domain (RBD) [15]. However, the omicron subvariant BF.7 carries an additional specific mutation, R346T in the spike protein of SARS-CoV-2 which is derived from the BA.4/5 subvariant, which is why the BF.7 variant has a 4.4-fold higher neutralization resistance than the original D614G variant [15]. The Arg346 mutations R346T in the BF.7 variant spike glycoprotein, particularly those on the RBD has been associated with an increased immune escape capability to neutralize antibodies generated by vaccines or previous infection [11-15]. A study by Wang, Qian et al., investigated the viral receptor affinities to dimeric human ACE2 by surface plasmon resonance and antibody evasion properties of the novel omicron subvariants BA.2, BA.4/5, BA.4.6, BA.4.7, BA.5.9, and BF.7. Findings of the study have shown that all the spike proteins from BA.4/5 sublineages, including BF.7 have shown a higher affinity for human ACE2[16].

### Effect of monoclonal antibodies and Vaccines against BF.7

The sensitivity profile of novel omicron subvariants to neutralisation by a panel of 23 mAbs found that the neutralisation profiles of BF.7, BA.4.6, BA.4.7 and BA.5.9 was similar to BA.4/5 except for mAbs in RBD class 3 (Bebtelovimab, Sotrovimab, romlusevimab, XGv289, XGv282, 2-7,ciglavimab, BD55-5840, BD55-3152,BD-804) which showed a substantial reduction in their neutralisation potency due to R346T mutation, which is associated with eliminated or weakened hydrogen bonds or salt bridges, or both, between R346 and some RBD class 3 mAbs [16]. A monoclonal antibody cocktail combination of cilgavimab and tixagevimab, has received an emergency use authorisation for the prevention of COVID-19, could also not neutralise BF.7 and other novel variants BA.4.6, BA.4.7, and BA.5.9[16].

A study by Sullivan, David J et al., to evaluate polyclonal antibodies from individuals both with at least 3 vaccine doses and also recently recovered from Omicron COVID-19(VaxCCP) neutralizing activity against new Omicron lineages in 740 individual patient samples from 37 separate cohorts defined by boosted vaccinations with or without recent Omicron COVID-19, as well as infection without vaccination found that more than 96% of the plasma samples from individuals in the recently i.e., within 6 months boosted VaxCCP study cohorts neutralized BQ.1.1 with 79%, XBB.1 with 22% and BF.7 with 94% variants respectively [17].

Another study by Sullivan DJ et al. found that the BA.5-bivalent booster dose vaccines elicited better neutralization against the newly emerged Omicron sublineages including BF.7 variant than the parental mRNA vaccine and those individuals with SARS-CoV-2 infection history develop higher and broader neutralization antibodies against the ongoing novel Omicron sublineages after the BA.5-bivalent booster [18].

Another study by Zhu, Ka-Li Jiang et al., found that two-dose CoronaVac or a third-dose ZF2001 booster elicits limited neutralization against Omicron subvariants 6 months after vaccination but hybrid immunity as well as Delta, BA.1, and BA.2 breakthrough infections induced long-term persistence of the antibody response, and over 70% of sera neutralized BF.7. as well as BA.1, BA.2, BA.4/BA.5[19].

In an in-vitro study by Wang, Qian et al., to investigate the antibody evasion properties of BA.4.6, BA.4.7, BA.5.9, and BF.7, we assessed the sensitivity of their corresponding pseudoviruses to neutralisation with serum samples from healthy individuals who had received three doses of a COVID-19 mRNA vaccine BNT162b2 or mRNA-1273 and patients with either BA.1, BA.2, or BA.4/5 breakthrough infection after vaccination found that the 50% inhibitory dose (ID_50_) titres of the booster dose vaccinated serum samples against BA.4.6, BA.4.7, BA.5.9, and BF.7 were similar to that against BA.4/5, with no more than 1·5-fold deviation in the geometric mean values and neutralizing antibody titres against new emerging omicron subvariants were higher than those of the serum samples of previous omicron variants[16].

Another systematic review of 27 included studies across the world on the effect of booster dose vaccination against Omicron and its subvariants by Chenchula S et al., found that booster-dose vaccines have produced significant efficacy against the SARS-CoV-2 and its subvariants [20].

## Discussion

The present review focuses on the variant of concern Omicron BF.7 with, increased binding to angiotensin-converting enzyme-2 (ACE2) receptor, increased expression of the RBD, enhanced infectivity and transmissibility potential with an increased immune evasion and resistance to the majority of available monoclonal antibodies is due to the R346T mutation in spike glycoprotein, particularly those on the RBD[15-16]. Being quicker to transmit than other Omicron variants, having an increased binding to angiotensin-converting enzyme-2 (ACE2) receptor, increased expression of the RBD, shorter incubation period, increased evasion of antibodies and a greater capacity to infect people who have had a previous COVID-19 infection, and/or been vaccinated, or both, the BF.7 variant is causing a significant increase in number COVI-19 cases in China and across the world [16]. A SARS-CoV-2 transmission model study by Kathy Leung et al., from China, using the data from recent outbreaks in Hong Kong and Shanghai to compare different scenarios in China concluded that the hospitals will be overwhelmed if infections rise as rapidly because of the easing of Zero COVID policy restrictions and will probably result in about one million deaths over the next few months, the study forecasts [21]. The study also suggests that, if 85% of the population gets a booster fourth dose of the COVID -19 vaccine other than the inactivated-virus vaccines, it could slow the rise in infections and reduce the number of severe infections and deaths by up to 35% [22]. According to this study, if China lifts the zero-COVID policy the Omicron could infect between 160 million and 280 million people and 1.55Lmillion deaths largely among unvaccinated older adults [22]. The study also suggests that, if 85% of the population gets the fourth dose of a vaccine other than the inactivated-virus vaccine could slow the increase in infections, the number of severe infections and deaths [22]. According to reports from China, more than 90% of the population has been fully vaccinated [23]. However, less than half of people aged 80 and over have received three doses of the vaccine [23]. Evidence also shows that the inactivated vaccine CoronaVac from China produced lower levels of neutralizing antibodies and these antibody levels also dropped quickly over time [23]. In addition, studies also have shown that protection from severe disease was also very less in preventing severe disease and death among 80 years and older [22]. Evidence from the literature suggests that the majority of monoclonal antibodies have not shown reduced neutralization potential against BF.7 variant. However, booster dose vaccination was significantly associated with increased efficacy against BF.7 and other emerging Omicron variants [16-20]. Therefore, the R346T mutation in the BF.7 variant was associated with reduced sensitivity to monoclonal antibodies, increased binding to angiotensin-converting enzyme-2 (ACE2) receptor, and increased expression of the RBD. Based on the evidence, COVID-19 infection with SARS-CoV-2 variant BF.7 may associate with a serious illness among the non-vaccinated population, with weaker immune systems such as the elderly and those with concomitant comorbidities [13-15].Limitations of the present review includes, there are only few number of studies available on BF.7 variant. Meta analysis was not done because of majority of studies are in vitro and there was no comparator.

## Conclusion

Findings of the present review suggest that infection with SARS-CoV-2 subvariant BF.7 with R346T mutation on the spike glycoprotein RBD, may be associated with increased transmissibility, and serious illness among the non-vaccinated and immune-compromised individuals such as the elderly, children and those with concomitant comorbidities. Therefore we recommend that, in addition to regular surveillance activities, the protection strategies need to rapidly achieve by global vaccination to nonvaccinated populations and booster dose vaccination with other than inactivated vaccines, especially among immunocompromised and vulnerable populations with comorbidities, those aged ≥60 years more and in children, to prevent possible outbreaks of novel SARS-CoV-2 Omicron subvariants and to decrease severity, mortality and to mitigate the healthcare and economic impacts due to the emerging Omicron sublineages and future VOCs.

## Data Availability

All data produced in the present study are available upon reasonable request to the authors

## Acknowledgement

None to declare.

## Conflict of Interests

The authors declare that there are no conflicts of interest.

## Grants and funding

None.

## Author Contributions

Santenna Chenchula conducted the literature search and data extraction and drafted the manuscript. Krishna Chaitanya Amarneni, Mohan Krishna Ghanta, Padmavathi R, Madhu Bhargavi Chandra, Madhu Babu Adusumilli, Bhawna Lakhawat, Madhavrao Chavan, Sofia Mudda, Rupesh Gupta revised the final manuscript. All authors reviewed and approved the final version of the manuscript.

## Data Availability

The data used in this systematic review is available from the corresponding author with a reasonable request.

## Notes

Sources of Support: Nil

Conflicts of interest: Nil

### Competing Interest Statement

The authors have declared no competing interest.

### Summary of Updates

Author names corrected and corrected grammatical mistakes.

